# Vasculopathy of the small vessels is common in lacunar stroke - a 7T MRI study

**DOI:** 10.64898/2026.07.09.26357711

**Authors:** Davor Pavlin-Premrl, Bradford Moffat, Rebecca Glarin, Vincent Thijs, Nawaf Yassi, Mark W Parsons, Peter J Mitchell, Julian Maingard, Hamed Asadi, Ashu Jhamb, Mark Schembri, Ali Khabaza, Anna Balabanski, Bruce CV Campbell

**Author notes:** **Corresponding author:** Dr Davor Pavlin-Premrl.

## Abstract

**Background:** Lacunar stroke is a common and disabling cerebrovascular disease. Small-vessel vasculopathy is thought to be the most common underlying cause, but this has only been identified on histopathology. 7T MRI allows small vessels to be seen in vivo. This study aimed to investigate rates of small vessel vasculopathy in lacunar stroke using 7T MRI.

**Methods:** Patients with lacunar stroke at an Australian tertiary stroke centre were prospectively screened and recruited to the study. Patients underwent 7T MRI with T1, T2, time-of-flight (TOF), diffusion-weighted imaging (DWI) and susceptibility-weighted imaging (SWI) sequences. Images were interpreted by two blinded neuroradiologists.

**Results:** The likely symptomatic perforator could be identified in 16/19 (84%) of cases. Amongst cases where the symptomatic perforator was observed, 14/16 (88%) of the symptomatic perforator vessels had focal stenosis consistent with steno-occlusive vasculopathy. There were 3/19 (16%) of cases with associated large artery vasculopathy. There were 7/16 (44%) cases where an occluded perforator was seen. The majority of patients had at least one vascular risk factor (15/19, 79%) and there were no cases where non-atherosclerotic vasculopathy was suspected.

**Conclusions:** Lacunar stroke is commonly associated with small vessel vasculopathy, likely due to atherosclerosis, which can be identified in vivo with 7T MRI time-of-flight imaging.

## Introduction

Lacunar stroke is a common and serious cerebrovascular disease, accounting for 15-26% of ischaemic stroke.^1^ It is associated with a ~20% recurrence rate and ~25% 5-year mortality.^2^

Lacunar strokes are due to occlusion of the small vessels, otherwise known as perforators.^3^ Occlusion of perforators associated with lacunar stroke has been observed in autopsies and brain imaging.^4^ Since Fisher’s seminal autopsy studies in the 1960s, it has been thought that the cause of these occlusions is small-vessel vasculopathy. Perforators ~300 to 800 μm in size are affected by atherosclerosis/arteriosclerosis similar to that seen in larger blood vessels, while perforators ~40 to 300 μm are affected by lipohyalinosis – characterised by fibrosis and accumulation of proteins and lipids.^5^ There has also been evidence published that lacunar stroke may be due to large-artery atherosclerosis at the origin of perforators^6^, and it has been argued that proximal embolic sources from AF or carotid atherosclerosis may be the cause of some perforator occlusions.^7^

7T MRI allows the small vessels implicated in lacunar stroke to be visualised. The higher field strength provides improved signal-to-noise ratio, and the longer tissue T1 at 7T enhances background suppression in TOF-MRA, together allowing perforators to be visualised non-invasively.^6^ 7T MRI has previously been used to investigate lacunar stroke, with studies showing that lacunar stroke is frequently associated with occluded perforator vessels^8^, occluded perforators can be seen to recanalise on repeat imaging^7^ and that lacunar stroke is frequently associated with ICAD at orifices of lenticulostriate arteries.^6^

The purpose of this study is to characterise the proportion of lacunar stroke that is associated with perforator vasculopathy. We hypothesised that the majority of lacunar stroke is associated with small-vessel vasculopathy visible on 7T MRI.

## Methods

Consecutive patients presenting to an Australian tertiary stroke centre from 2016-2020 were prospectively screened for acute lacunar stroke using MRI results. Patients with concurrent cortical stroke were excluded. Those who were clinically well enough to undergo transfer to the neighbouring research institution and who consented to the study were enrolled.

Clinical data were collected, including: demographic data; modified Rankin Scale (mRS); vascular risk factors; presence of possible proximal embolic source; evidence of non-atherosclerotic intracranial vasculopathy such as vasculitis, dissection, infective vasculopathy or genetic vasculopathy; National Institutes of Health Stroke Scale (NIHSS); presence of stuttering clinical course consistent with capsular warning syndrome and whether thrombolysis was administered.

MRI scans were performed at the Melbourne Brain Centre Imaging Unit on a 7-Tesla Siemens MAGNETOM Terra scanner. The protocol utilised a 32-channel receive / 1-channel transmit head coil (Nova Medical). The sequences obtained were T1 Magnetization-Prepared 2 Rapid Acquisition Gradient Echo (MP2RAGE), 3D T2-weighted Sampling Perfection with Application-Optimized Contrasts using Different Flip Angle Evolution (SPACE), Susceptibility-Weighted Imaging (SWI), Diffusion-Weighted Imaging (DWI) and Time-of-Flight (TOF). A limited series of unmatched healthy control scans was obtained from study investigators.

Imaging interpretation was performed by neuroradiologists and an interventional neuroradiologist. Assessment of perforators was performed by two blinded neuroradiologists, with a third interventional neuroradiologist arbitrating any cases of disagreement. All interpreters have more than 10 years experience. Imaging data from 7T MRI as well as other available imaging was reviewed. The degree of artefact was evaluated using a Likert scale^9^ of 1 to 4: 1) images contain severe artefact and cannot be used for diagnostic purposes, 2) images contain significant artefacts but can be used for gross diagnostic purposes, 3) images contain some artefacts but can be used for diagnostic purposes and 4) images do not contain artefacts. Lacunar infarcts were identified on DWI. White matter hyperintensities were identified on FLAIR sequences and assessed using the Fazekas rating system.^10^ Microbleeds and superficial siderosis were identified on SWI sequences. The Microbleed Anatomical Rating Scale (MARS) was used.^11^ Time-of-flight sequences were used to identify large artery vasculopathy, the likely symptomatic perforator, small-vessel vasculopathy and perforator occlusion. Large-vessel vasculopathy was evaluated using the Warfarin-Aspirin Symptomatic Intracranial Disease (WASID) method, where the vessel proximal to the stenosis is considered the baseline normal segment. Small-vessel vasculopathy was defined by the presence of stenosis or by the presence of signal loss and then re-emergence. Severity of small vessel steno-occlusive disease was graded as severe (near-occlusion) or mild-moderate. The likely symptomatic small vessel was also evaluated for the presence of vessel occlusion. The infarcts were classified as anterior or posterior circulation based on the location of the infarct.

Statistical analysis was limited to descriptive statistics due to low patient numbers.

## Results

Our study recruited 19 patients. The mean age was 64 and 58% of patients had hypertension. No patients had clinical evidence of non-atherosclerotic vasculopathy. The demographic and clinical data of patients is summarised in table 1.

**Table 1.** patient characteristics.

|  |  |
| --- | --- |
| Age (mean) | 64 (SD±10.1) |
| Female % | 5/19 (26%) |
| Any vascular risk factor (%) | 15/19 (79%) |
| Hypertension (%) | 11/19 (58%) |
| Diabetes mellitus (%) | 4/19 (21%) |
| Dyslipidaemia (%) | 5/19 (26%) |
| Atrial fibrillation (%) | 2/19 (11%) |
| Smoking (%) | 5/19 (26%) |
| Previous stroke/TIA (%) | 0/19 (0%) |
| Evidence of non-atherosclerotic vasculopathy | 0/19 (0%) |
| Baseline NIHSS (median) | 3 |
| Baseline mRS (median) | 0 |
| Possible embolic source (cardioembolic, proximal atherosclerosis) | 4/19 (21%) |
| Thrombolysis given | 3/19 (16%) |
| History of capsular warning syndrome | 1/19 (5%) |

Likely symptomatic perforator could be identified in 16/19 cases (84%). For anterior circulation lacunar strokes, the symptomatic perforator could be observed in 7/8 (88%) of cases. For posterior circulation lacunar stroke, the symptomatic perforator could be observed in 9/11 (82%) of cases. Two of the three cases where the symptomatic perforator was not identified were brainstem strokes. Brainstem perforators were not visible on any of the 7T scans performed due to technical limitations of low signal in that region. Amongst cases where the symptomatic perforator was observed, 14/16 (88%) of the symptomatic perforator vessels had steno-occlusive vasculopathy. There were 3/19 (16%) of cases with associated large artery vasculopathy. There were 7/16 (44%) cases where an occluded perforator was seen. Imaging findings are summarised in table 2.

**Table 2.**
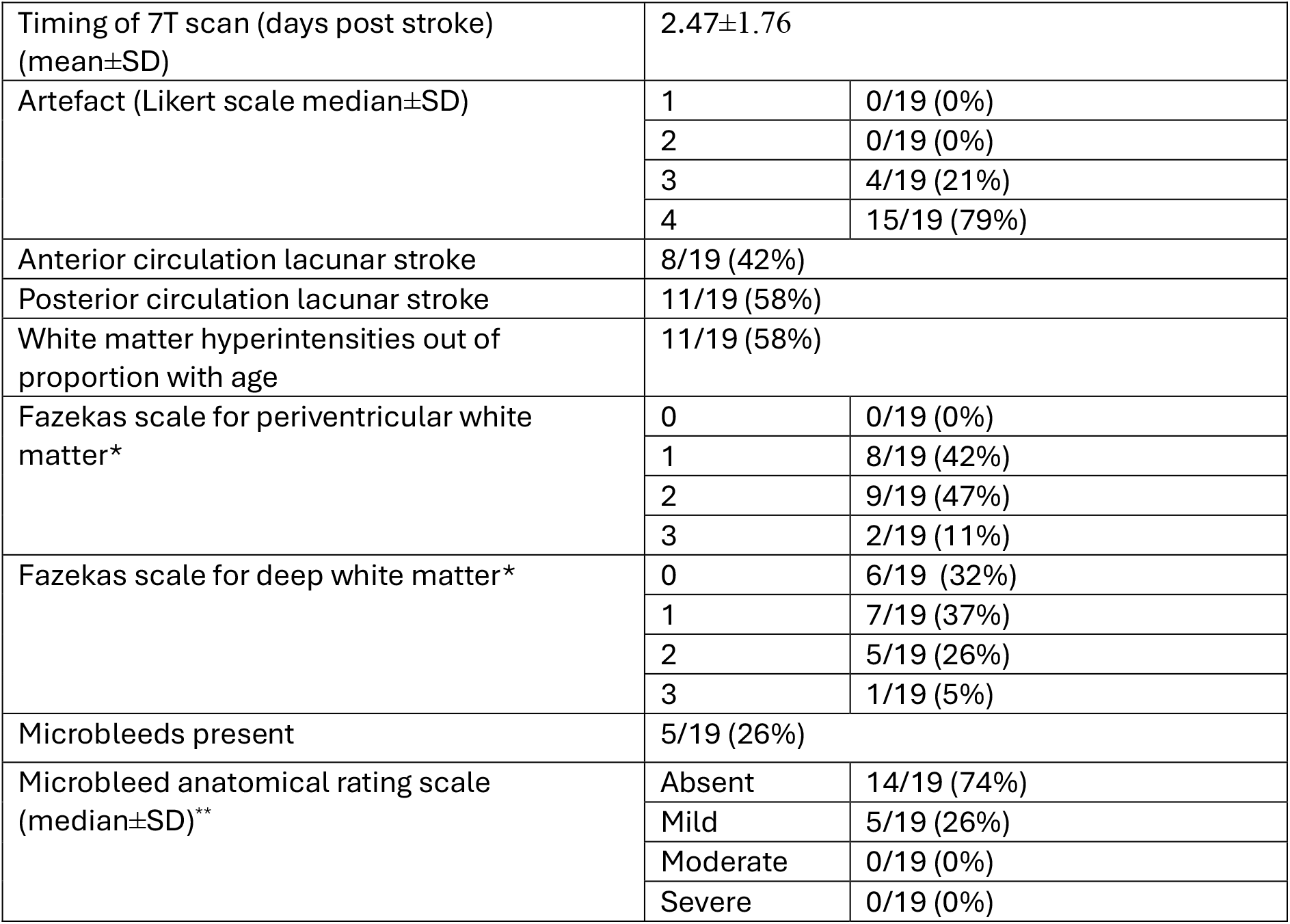
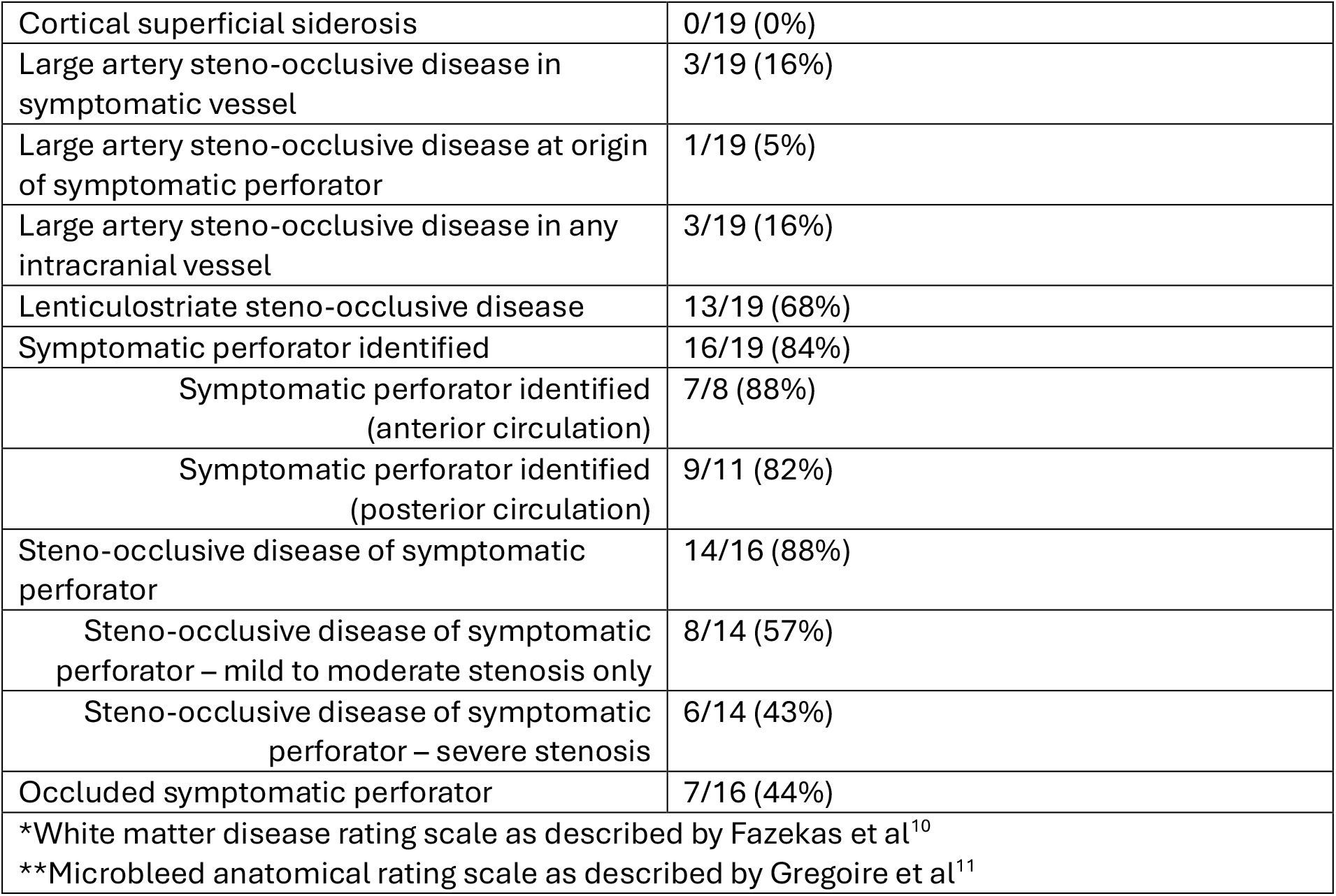
imaging findings.

|  |  |  |
| --- | --- | --- |
| Timing of 7T scan (days post stroke) (mean±SD) | 2.47±1.76 |  |
| Artefact (Likert scale median±SD) | 1 | 0/19 (0%) |
|  | 2 | 0/19 (0%) |
|  | 3 | 4/19 (21%) |
|  | 4 | 15/19 (79%) |
| Anterior circulation lacunar stroke | 8/19 (42%) |  |
| Posterior circulation lacunar stroke | 11/19 (58%) |  |
| White matter hyperintensities out of proportion with age | 11/19 (58%) |  |
| Fazekas scale for periventricular white matter* | 0 | 0/19 (0%) |
|  | 1 | 8/19 (42%) |
|  | 2 | 9/19 (47%) |
|  | 3 | 2/19 (11%) |
| Fazekas scale for deep white matter* | 0 | 6/19 (32%) |
|  | 1 | 7/19 (37%) |
|  | 2 | 5/19 (26%) |
|  | 3 | 1/19 (5%) |
| Microbleeds present | 5/19 (26%) |  |
| Microbleed anatomical rating scale (median±SD)** | Absent | 14/19 (74%) |
|  | Mild | 5/19 (26%) |
|  | Moderate | 0/19 (0%) |
|  | Severe | 0/19 (0%) |

|  |  |
| --- | --- |
| Cortical superficial siderosis | 0/19 (0%) |
| Large artery steno-occlusive disease in symptomatic vessel | 3/19 (16%) |
| Large artery steno-occlusive disease at origin of symptomatic perforator | 1/19 (5%) |
| Large artery steno-occlusive disease in any intracranial vessel | 3/19 (16%) |
| Lenticulostriate steno-occlusive disease | 13/19 (68%) |
| Symptomatic perforator identified | 16/19 (84%) |
| Symptomatic perforator identified (anterior circulation) | 7/8 (88%) |
| Symptomatic perforator identified (posterior circulation) | 9/11 (82%) |
| Steno-occlusive disease of symptomatic perforator | 14/16 (88%) |
| Steno-occlusive disease of symptomatic perforator – mild to moderate stenosis only | 8/14 (57%) |
| Steno-occlusive disease of symptomatic perforator – severe stenosis | 6/14 (43%) |
| Occluded symptomatic perforator | 7/16 (44%) |
| *White matter disease rating scale as described by Fazekas et al<sup>10</sup> |  |
| **Microbleed anatomical rating scale as described by Gregoire et al<sup>11</sup> |  |

Seven healthy control scans were obtained from study investigators, with a mean age of 35.5. No controls had any vascular risk factors or medical history. No control scans demonstrated stenosis of small vessels or large vessels, infarcts, white matter disease or evidence of amyloid angiopathy.

## Discussion

This study demonstrates that steno-occlusive disease of lenticulostriates is common in lacunar stroke and can be demonstrated on 7T MRI. Lenticulostriate vasculopathy was present in the majority of symptomatic perforators (14/16, 88%). Widespread lenticulostriate steno-occlusive disease was also present in the majority of cases (13/19, 68%). Given the mean age, presence of vascular risk factors and absence of non-atherosclerotic etiologies, the cause of steno-occlusive disease in our cohort is likely small vessel atherosclerosis or lipohyalinosis. In contrast to the small vessels, concurrent large artery vasculopathy was present in a minority of cases. Steno-occlusive disease in the symptomatic large vessel was seen in 3/19 (16%) cases and disease at the origin of perforators seen in 1/19 (5%). The discordance between rates of large artery atherosclerosis and small vessel atherosclerosis suggests that lenticulostriate vessels are particularly vulnerable to vascular risk factors, possibly due to their small size relative to the feeding arteries. Capsular warning syndrome, where lacunar stroke symptoms have a stuttering course, is thought to be due to lenticulostriate atherosclerosis causing haemodynamic insufficiency. Despite the majority of symptomatic perforators in our study having steno-occlusive disease, only 1/19 (5%) of cases had capsular warning syndrome. This suggests that absence of a stuttering time course does not imply absence of underlying vasculopathy.

It is hypothesized that lacunar stroke is due to small-vessel occlusion. In this study, vessel occlusion in the symptomatic perforator was identified in approximately half of cases (7/16, 44%). This may be due to spontaneous recanalization, which has previously been described in 7T studies in lacunar stroke and is observed in patients with occlusion of larger vessels.^7^ It may also be due to the occlusion being below the resolution of 7T MRI. Another possible explanation is that lacunar stroke in the setting of small vessel atherosclerosis may occur due to haemodynamic insufficiency without vessel occlusion, as is seen with large artery atherosclerosis.

Anterior circulation perforator vessels were readily visible on 7T MRI with the symptomatic perforator identified in all but one case. In the posterior circulation, thalamic perforators were visible but not brainstem perforators, resulting in the symptomatic perforator being identified in 9/11 (82%) of cases.

This study supports the hypothesis that lacunar stroke should be thought of as primarily a disease of small vessel vasculopathy, likely secondary to atherosclerosis. Medical therapy for large artery atherosclerosis has improved significantly in recent decades, with dual antiplatelet therapy (DAPT) and aggressive risk factor control becoming standard of care.^12^ Similar strategies are likely important in lacunar stroke. Although the SPS3 trial did not show a benefit with dual antiplatelet therapy, this was likely due to the delay in commencement and length of therapy.^13^ Dual antiplatelet therapy in lacunar stroke is supported by a sub-analysis of the Cilostazol Stroke Prevention Study (CSPS) studies, but further lacunar-specific trials are needed.^14^ Novel therapies currently being explored in large artery atherosclerosis such as lipid-lowering with PCSK9 inhibitors^15^ and treatment of the inflammatory cascade^12^ could be explored in lacunar stroke. 7T MRI could play an important role in investigation of lacunar stroke therapies as the only non-invasive method of imaging perforator vessels. The technology could be used to identify diseased vessels and to monitor changes with therapies.

Our study had a number of limitations, the most significant being small patient numbers, limiting the generalisability of results. Although our study had control patients, these were not matched or equal in number to the lacunar stroke cases. There are also some inherent limitations to 7T MRI. The technology is very sensitive to movement artefact which limited interpretation of some scans.

## Conclusion

7T MRI can be used to identify atherosclerosis of lenticulostriates. Our study supports the hypothesis that lacunar stroke is due to small-vessel atherosclerosis.

## Data Availability

Data is available on reasonable request

## Ethics statement

The study protocol was reviewed and approved by the Melbourne Health Human Research Ethics Committee (2013.299).

---

**Figure 1.**
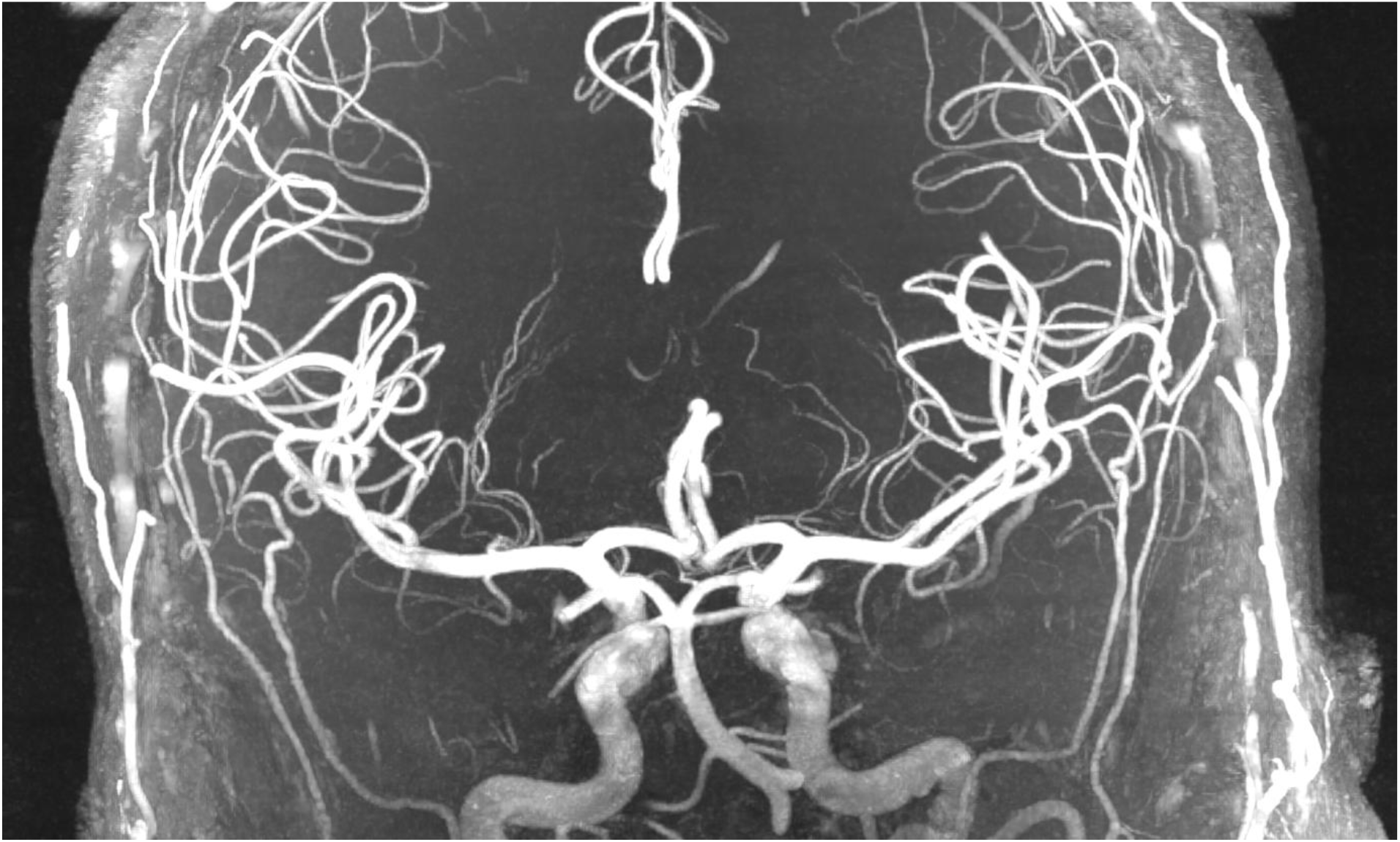
7T TOF in healthy control. Lenticulostriates are well visualised, contiguous with no abrupt occlusions.

**Figure 2.**
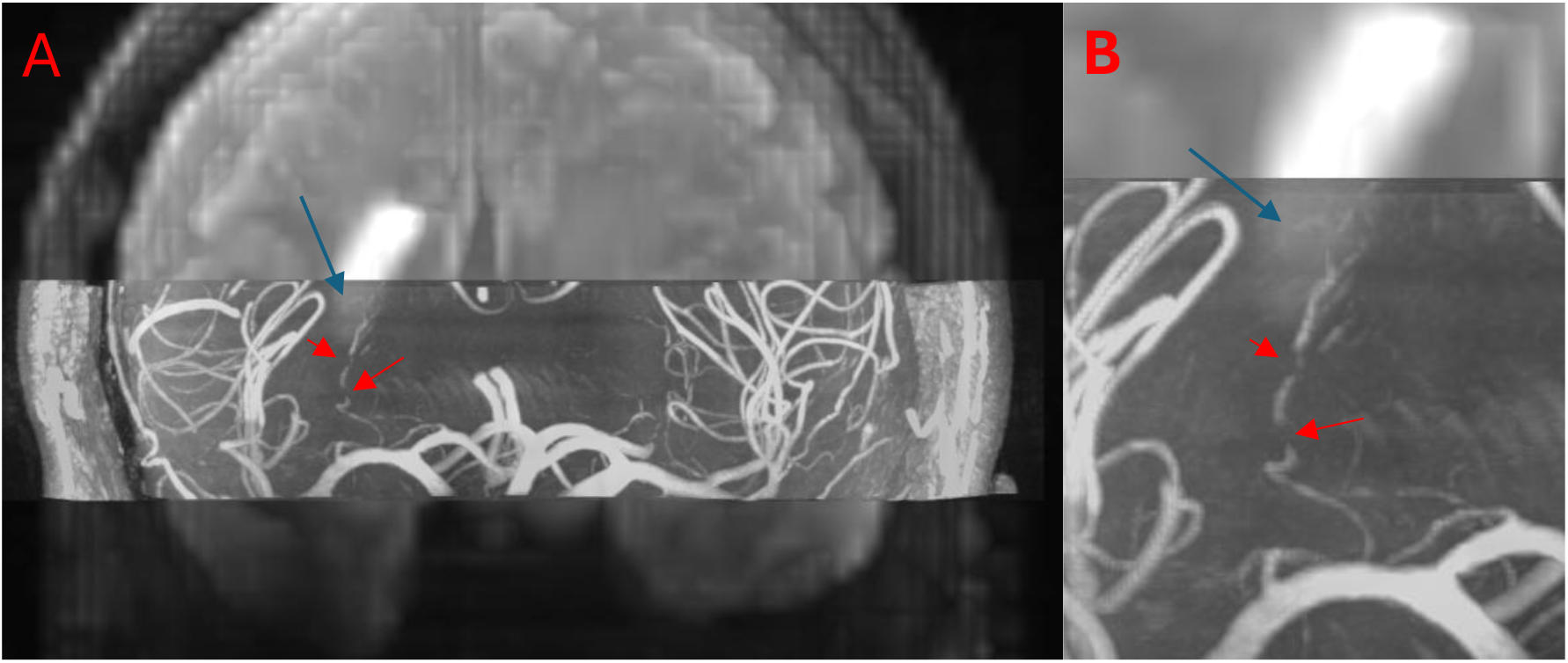
Overlay of 7T MRI DWI and TOF sequences (note TOF sequence has a narrower slab than the DWI). Right lacunar stroke seen on DWI sequence (blue arrow). The symptomatic perforator is affected by severe, multi-focal atherosclerosis (red arrows), seen on whole-brain (A) and magnified (B) views.

**Figure 3.**
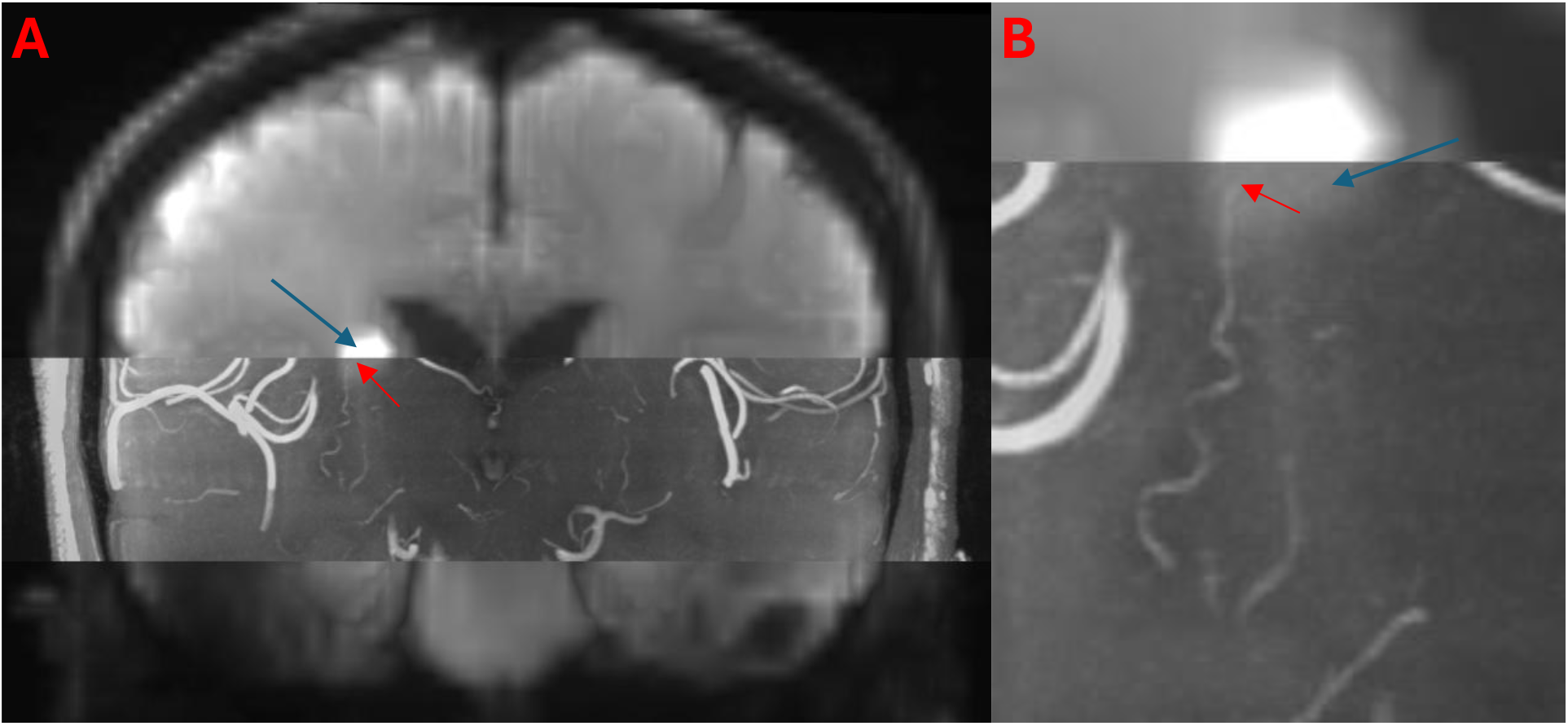
Overlay of 7T MRI DWI and TOF sequences (note TOF sequence has a narrower slab than the DWI). Right lacunar infarct labelled with blue arrow. Vessel cut-off (red arrows) seen on whole-brain (A) and magnified (B) views.

**Figure 4.**
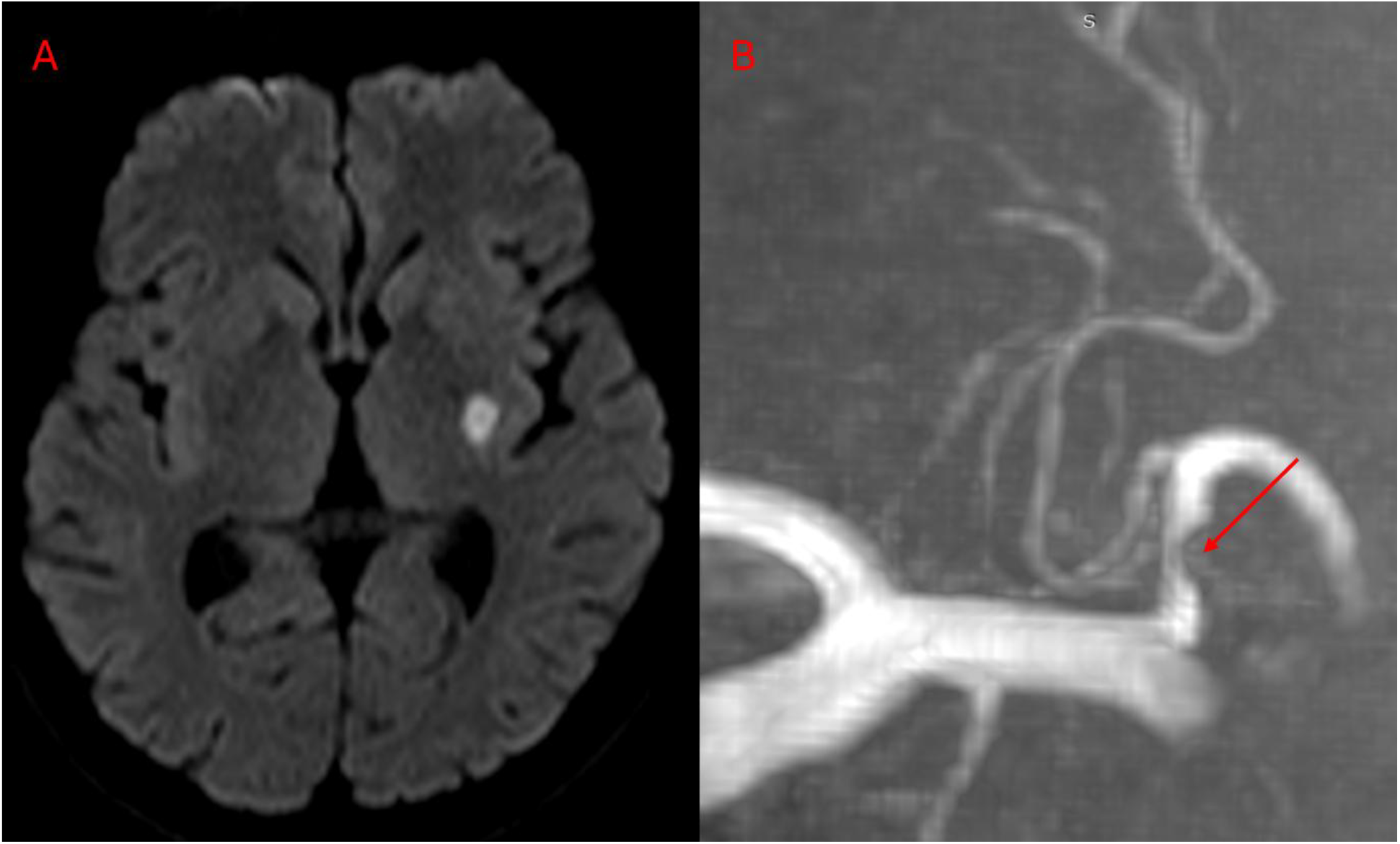
This patient had left posterior limb of internal capsule infarct (A). All visible lenticulostriate vessels branch from the M2 segment, rather than the typical M1 segment (B). The M2 segment has intracranial atherosclerosis proximal to the lenticulostriate take-off (arrow). This case demonstrates how 7T MRI can assist in understanding the pathophysiology of particular lacunar stroke patients, as without visualising the lenticulostriate vessels it would be assumed that large vessel atherosclerosis had not contributed to this patient’s stroke.

